# Spinal cord MRI and MRS Detect Early-stage Alterations and Disease Progression in Friedreich Ataxia

**DOI:** 10.1101/2022.01.28.22270048

**Authors:** James M. Joers, Isaac M. Adanyeguh, Dinesh K. Deelchand, Diane H. Hutter, Lynn E. Eberly, Isabelle Iltis, Khalaf O. Bushara, Christophe Lenglet, Pierre-Gilles Henry

## Abstract

**Background:** Friedreich Ataxia (FRDA) is the most common hereditary ataxia. Atrophy of the spinal cord is one of the hallmarks of the disease. Magnetic resonance imaging (MRI) and spectroscopy (MRS) are powerful and non-invasive tools to investigate pathological changes in the spinal cord. A handful of studies have reported *cross-sectional* alterations in FRDA using MRI and diffusion MRI (dMRI) in FRDA. However, to our knowledge no *longitudinal* MRI, dMRI or MRS results have been reported in the spinal cord in FRDA.

**Objective:** To investigate early-stage cross-sectional alterations and longitudinal changes in the cervical spinal cord in FRDA, using a multimodal magnetic resonance (MR) protocol comprising morphometric (anatomical MRI), microstructural (dMRI), and neurochemical (^1^H MRS) assessments.

**Design:** We enrolled 28 early-stage individuals with FRDA and 20 age- and gender-matched controls (cross-sectional study). Disease duration at baseline was 5.5±4.0 years and Friedreich Ataxia Rating Scale (FARS) total neurological score at baseline was 42.7±13.6. Twenty-one FRDA participants returned for 1-year follow-up, and 19 of those for 2-year follow-up (cohort study). Each visit consisted in clinical assessments and MR scans. Controls were scanned at baseline only.

**Results:** At baseline, individuals with FRDA had significantly lower spinal cord cross-sectional area (−31%, p=4.10^−17^), higher eccentricity (+10%, p=5.10^−7^), lower total N-acetyl-aspartate (−36%, p=6.10^−9^) and higher myo-inositol (+37%, p=2.10^−6^) corresponding to a lower ratio tNAA/mIns (−52%, p=2.10^−13^), lower fractional anisotropy (−24%, p=10^−9^) as well as higher radial diffusivity (+56%, p=2.10^−9^), mean diffusivity (+35%, p=10^−8^) and axial diffusivity (+17%, p=4.10^−5^) relative to controls.

Longitudinally, spinal cord cross-sectional area decreased by 2.4% per year relative to baseline (p=4.10^−4^), the ratio tNAA/mIns decreased by 5.8% per year (p=0.03), and fractional anisotropy showed a trend to decrease (−3.2% per year, p=0.08).

Spinal cord cross-sectional area correlated strongly with clinical measures, with the strongest correlation coefficients found between cross-sectional area and Scale for the Assessment and Rating of Ataxia (SARA) (R=-0.55, p=7.10^−6^) and between cross-sectional area and FARS total neurological score (R=-0.60, p=4.10^−7^). Less strong but still significant correlations were found for fractional anisotropy and tNAA/mIns.

**Conclusion:** We report here the first quantitative longitudinal MR results in the spinal cord in FRDA. The largest longitudinal effect size was found for spinal cord cross-sectional area, followed by tNAA/mIns and fractional anisotropy. Our results provide direct evidence that abnormalities in the spinal cord result not solely from hypoplasia, but also from neurodegeneration, and show that disease progression can be monitored non-invasively in the spinal cord.

## INTRODUCTION

Friedreich ataxia (FRDA) is the most common inherited ataxia, with an incidence ranging from 1:20,000 to 1:250,000, or less depending on countries ^1,2^. Genetic transmission is autosomal recessive and is caused by mutations in the frataxin gene (in most cases a GAA repeat expansion in intron 1) leading to reduced expression of frataxin ^3,4^. Symptoms include limb and gait ataxia, dysarthria and cardiomyopathy ^5^. Onset is typically in the second decade of life ^6^ and there is currently no effective disease-modifying treatment.

Nikolaus Friedreich recognized spinal cord degeneration as a hallmark of the disease ^7^ and spinal cord atrophy was reported in early MRI studies ^8-12^. In the pathogenesis of FRDA, substantial demyelination and gliosis of the posterior and lateral columns of the spinal cord occur prior to cerebellar pathology ^5^. In particular, dorsal root ganglia, posterior roots and posterior columns of the spinal cord show significant pathological alterations ^13-15^. Recent cross-sectional MRI studies showed that spinal cord cross-sectional area (CSA) is significantly smaller in FRDA than in controls and correlates negatively with disease severity ^15-17^. However, longitudinal MR data documenting disease progression in the spinal cord are still lacking. With many clinical trials ongoing or on the horizon, including gene therapy trials, MR could play a key role in assessing disease progression and the effect of prospective treatments. Consequently, “natural history” longitudinal MR data documenting disease progression in the spinal cord are urgently needed.

Beyond anatomical imaging, advanced MR modalities, such as diffusion MRI (dMRI) and magnetic resonance spectroscopy (MRS) have the potential to respectively reveal microstructural and neurochemical alterations not visible in conventional anatomical images.

Diffusion MRI ^18^ relies on the anisotropic diffusion of water in organized tissues, such as the brain white matter or spinal cord, to recover microstructural and connectivity information through local biophysical modeling and tractography ^19^. Axonal membranes hinder the diffusion of water molecules, a phenomenon that can be quantified through MRI by taking measurements along multiple orientations. Diffusion tensor imaging (DTI) models diffusion MRI data by assuming Gaussian diffusion at each location of the imaged tissue. Diffusion MRI of the spinal cord is challenging and has only recently received growing attention and seen new developments in the neuroimaging community ^20-26^, with applications in disorders such as degenerative cord compression ^27^, multiple sclerosis ^28^, amyotrophic lateral sclerosis ^23^ and traumatic spinal cord injury ^29^. To the best of our knowledge, only one study has reported cross-sectional data using dMRI in the spinal cord in FRDA ^30^, and no longitudinal dMRI data have been published.

Proton Magnetic Resonance Spectroscopy (^1^H-MRS) allows non-invasive measurement of the concentration of multiple metabolites in multiple organs, including the spinal cord. Although challenging to implement, ^1^H-MRS in the spinal cord has been shown to be feasible ^31-34^. A number of studies have reported neurochemical alterations in the spinal cord in neurodegenerative diseases (Blamire, Cader et al. 2007, Marliani, Clementi et al. 2010, Elliott, Pedler et al. 2011), and changes in N-acetyl-aspartate (NAA) concentrations in a longitudinal assessment of lesions of the spinal cord in multiple sclerosis (Ciccarelli, Altmann et al. 2010). However, no data have been reported using MRS in the spinal cord in FRDA.

The objective of the present study was to study pathological alterations and longitudinal changes in the cervical spinal cord in an early-stage cohort of individuals with FRDA, using an advanced multimodal MR protocol (MRI, dMRI and MRS). We hypothesized that FRDA participants would show significant alterations in morphometry, microstructure and neurochemistry relative to controls, even at early-stage, and that FRDA participants would show changes over time.

## MATERIALS AND METHODS

### Participants

All procedures were conducted in accordance with the Declaration of Helsinki and were approved by the University of Minnesota Institutional Review Board. All participants provided informed consent (adults) or, for minors, informed assent with their parents providing informed consent.

Twenty-eight individuals with FRDA (mean age 19.0 ± 7.3 years, range 11-35, 14M/14F) and 20 age- and gender-matched control individuals (20.4 ± 7.1 years, range 10-35, 11M/9F) were recruited for this study (Table 1). The FRDA group was at an early stage of the disease (Time from diagnosis: 2.3 ± 2.6 years; Disease duration: 5.5 ± 4.0 years). FRDA participants were recruited through the University of Minnesota Ataxia Research Center and the Friedreich’s Ataxia Research Alliance (FARA). Control participants were recruited through the local CMRR volunteer list and word of mouth. Eligibility criteria for FRDA participants were: confirmed FRDA diagnosis. Efforts were made to primarily enroll FRDA participants at an early-stage of the disease. Only two FRDA participants were wheelchair-bound at baseline. Exclusion criteria for controls were: neurological disease. Exclusion criteria for all participants were: contraindication for MR scan, pregnancy, claustrophobia, diabetes, smoking, restless leg syndrome, history of alcoholism. All subjects were asked to withhold antioxidants such as multivitamins for 3 weeks prior to scanning. All subjects were also asked to withhold benzodiazepine or any medication specific to ataxia symptoms for 36 hours prior to scanning.

**Table 1.**
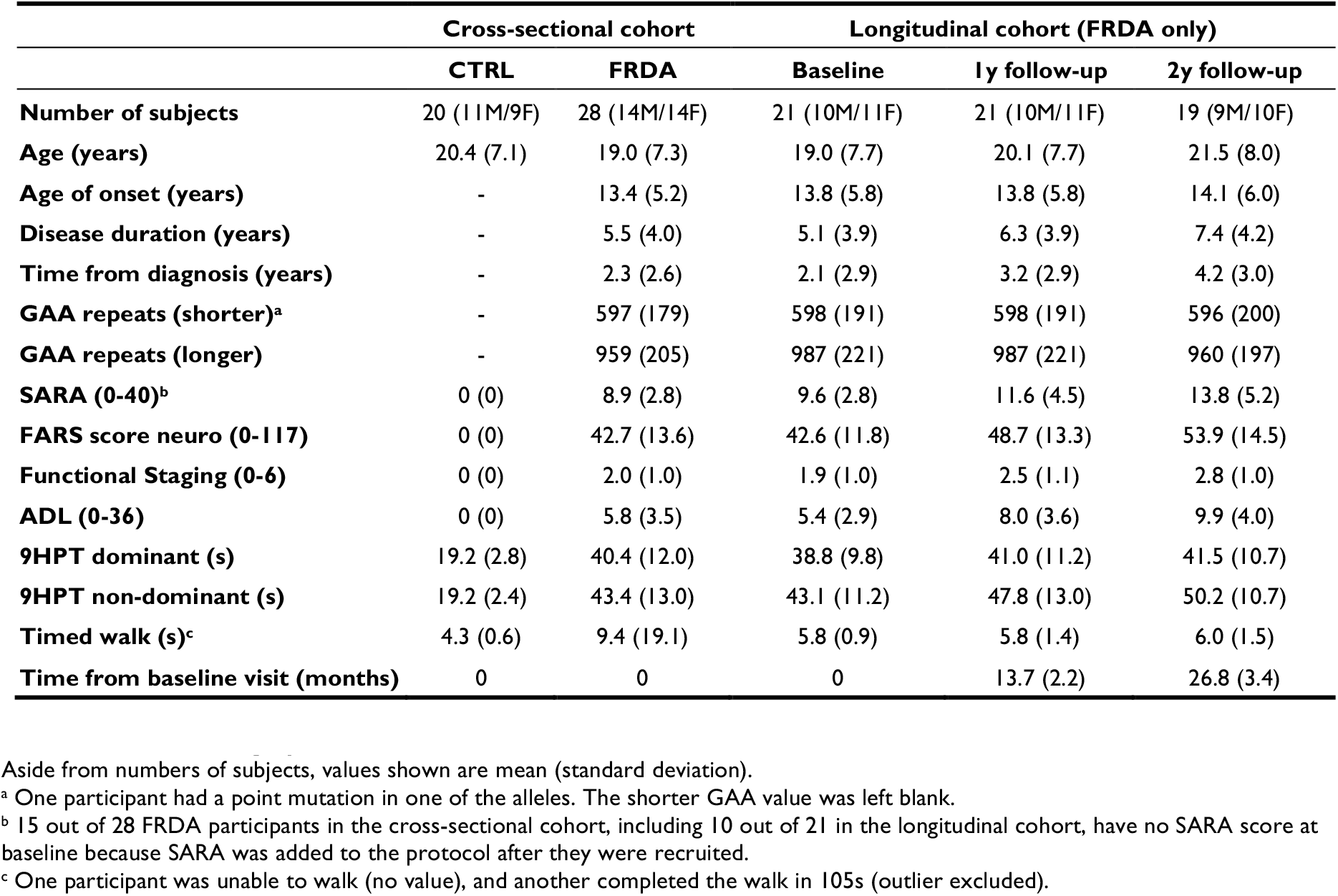
Cohort demographic and clinical characteristics at baseline.

|  | Cross-sectional cohort |  | Longitudinal cohort (FRDA only) |  |  |
| --- | --- | --- | --- | --- | --- |
|  | CTRL | FRDA | Baseline | 1y follow-up | 2y follow-up |
| **Number of subjects** | 20 (11M/9F) | 28 (14M/14F) | 21 (10M/11F) | 21 (10M/11F) | 19 (9M/10F) |
| **Age (years)** | 20.4 (7.1) | 19.0 (7.3) | 19.0 (7.7) | 20.1 (7.7) | 21.5 (8.0) |
| **Age of onset (years)** | - | 13.4 (5.2) | 13.8 (5.8) | 13.8 (5.8) | 14.1 (6.0) |
| **Disease duration (years)** | - | 5.5 (4.0) | 5.1 (3.9) | 6.3 (3.9) | 7.4 (4.2) |
| **Time from diagnosis (years)** | - | 2.3 (2.6) | 2.1 (2.9) | 3.2 (2.9) | 4.2 (3.0) |
| **GAA repeats (shorter)<sup>a</sup>** | - | 597 (179) | 598 (191) | 598 (191) | 596 (200) |
| **GAA repeats (longer)** | - | 959 (205) | 987 (221) | 987 (221) | 960 (197) |
| **SARA (0-40)<sup>b</sup>** | 0 (0) | 8.9 (2.8) | 9.6 (2.8) | 11.6 (4.5) | 13.8 (5.2) |
| **FARS score neuro (0-117)** | 0 (0) | 42.7 (13.6) | 42.6 (11.8) | 48.7 (13.3) | 53.9 (14.5) |
| **Functional Staging (0-6)** | 0 (0) | 2.0 (1.0) | 1.9 (1.0) | 2.5 (1.1) | 2.8 (1.0) |
| **ADL (0-36)** | 0 (0) | 5.8 (3.5) | 5.4 (2.9) | 8.0 (3.6) | 9.9 (4.0) |
| **9HPT dominant (s)** | 19.2 (2.8) | 40.4 (12.0) | 38.8 (9.8) | 41.0 (11.2) | 41.5 (10.7) |
| **9HPT non-dominant (s)** | 19.2 (2.4) | 43.4 (13.0) | 43.1 (11.2) | 47.8 (13.0) | 50.2 (10.7) |
| **Timed walk (s)<sup>c</sup>** | 4.3 (0.6) | 9.4 (19.1) | 5.8 (0.9) | 5.8 (1.4) | 6.0 (1.5) |
| **Time from baseline visit (months)** | 0 | 0 | 0 | 13.7 (2.2) | 26.8 (3.4) |

Aside from numbers of subjects, values shown are mean (standard deviation).
<sup>a</sup> One participant had a point mutation in one of the alleles. The shorter GAA value was left blank.
<sup>b</sup> 15 out of 28 FRDA participants in the cross-sectional cohort, including 10 out of 21 in the longitudinal cohort, have no SARA score at baseline because SARA was added to the protocol after they were recruited.
<sup>c</sup> One participant was unable to walk (no value), and another completed the walk in 105s (outlier excluded).

Twenty-one participants with FRDA returned for follow-up (referred to as the “longitudinal cohort”) approximately one year later (time between baseline and 1-year follow-up = 13.7 ± 2.2 months), and 19 participants also returned for a 2-year follow-up (time between baseline and 2-year follow-up = 26.8 ± 3.4 months). In the following, we refer to “1-year” and “2-year” follow-ups for brevity. Controls were scanned only at baseline. Reasons for not returning for 1-year and 2-year follow-up included: too progressed (1), rod placement (2), brain arteriovenous anomaly (1), claustrophobia (1) and no longer interested or too busy (4).

Study visits took place at the Center for Magnetic Resonance Research, University of Minnesota from 2013 to 2018. Each visit included three ∼1hr scanning sessions at 3 Tesla over 1-2 days: One for brain anatomical and diffusion MRI, one for spinal cord dMRI, and one for spinal cord MRS. Because of the large amount of data, we focus in this paper on the spinal cord findings, with the brain anatomical MRI used only for cervical spinal cord morphometry.

In addition to MR scans, all patients were assessed with the original Friedreich Ataxia Rating Scale (FARS) ^35^, which includes total neurological score (FARS total neuro, range 0-117), activities of daily living (ADL, range 0-36), functional staging (range 0-6), timed 9-hole peg test (9HPT) with dominant and non-dominant hand and timed 25ft walk test. In addition, Scale for the Assessment and Rating of Ataxia (SARA) scores (range 0-40) were collected except for 15 FRDA participants at baseline.

### MR Data Acquisition

Measurements were initially performed on a 3 Tesla Siemens MAGNETOM Trio scanner (Siemens, Erlangen, Germany) with Syngo software version VD13D. The Trio scanner was upgraded to a MAGNETOM Prisma^fit^ scanner with Syngo software version VE11C during the study. The potential impact of this upgrade was carefully investigated and additional scans and analyses were performed to ensure that the upgrade did not bias our results (see “Scanner upgrade” section below).

On both scanners, the body transmit RF coil was used. On Trio, a 32-channel receive head coil was used for brain acquisitions, and a 20-channel receive head-neck coil was used for spine acquisitions. On Prisma, a 64-channel receive head-neck coil was used for all acquisitions.

#### Brain Anatomical MRI

Brain coronal T_1_-weighted 3D MPRAGE images were obtained with the following parameters: voxel size 1mm isotropic, TR/TE=2530/3.65ms, TI=1100ms, flip angle= 7°, 224 slices, field of view = 256×176 mm^2^, phase encode in R-L direction, 2x GRAPPA.

#### Spinal Cord Diffusion MRI

Sagittal T_2_-weighted 2D TSE images with low resolution in the slice direction were acquired for positioning the dMRI volume of interest and for identification of cervical levels, with the following parameters: field of view 260×260 mm^2^, matrix 384×384, 13 slices, 3mm slice thickness, 0.7×0.7 mm^2^ in-plane resolution, TR/TE=3500/112ms, flip angle 160deg.

The cervical spine was scanned axially, covering levels C2 to C7, with 1.12×1.12 mm^2^ (field of view 118×62 mm^2^) in-plane resolution and 3.3mm slice thickness (30 slices), as illustrated in Figure 2. A segmented-readout EPI sequence (RESOLVE) ^36^ was used in combination with parallel imaging to reduce artifacts due to susceptibility changes at tissue interfaces around the spine, as previously reported. A finger pulse oximeter was used for cardiac triggering. The following parameters were used: TR/TE=4500/66ms, in-plane acceleration (iPAT) = 2, 30 directions, *b*-value 650 s/mm^2^, six *b*=0 volumes. Data were collected with reversed phase-encode blips (AP and PA), resulting in pairs of images with distortions going in opposite directions. From these pairs the susceptibility-induced off-resonance field was estimated and corrected using a method similar to that described in ^37^. Subject motion and eddy-current induced distortions were corrected ^38^ as previously reported ^23^.

#### Spinal Cord Magnetic Resonance Spectroscopy

Sagittal T_2_-weighted images similar to those acquired before spinal cord diffusion were used to position an 8×6×30 mm^3^ (1.44 ml) voxel in the spine, centered on the C4-C5 intervertebral disc. B_0_ shimming was performed using Siemens image-based shimming with a reduced field of view. Average water linewidth in the voxel was 13.0 ± 3.4 Hz after shimming.

MR spectra were acquired using an optimized semi-LASER sequence with TE=28ms, TR=5s, and 2 × 128 averages. A detailed description of the sequence, including outer-volume suppression and VAPOR water suppression can be found in ^39^. Individual shots were saved separately. A finger pulse oximeter was used for cardiac triggering. Unsuppressed water reference signals (NT=4) were collected for eddy-current correction, and to serve as an internal concentration reference for quantification. Water signal was also measured at different TEs (28, 35, 50, 70, 100, 200, 300,500, 800, 1000, 1500, 3000 and 4000ms) with one average per TE and TR=15sec. The resulting T_2_ curve was fit with a bi-exponential function to determine the fraction of CSF and tissue in the voxel, assuming a T_2_ value of 740ms for CSF. The entire procedure was repeated twice, yielding two spectra with 128 transients each, for a total of 256 transients.

Both individual shots and accumulating spectrum were monitored in real time for spectral quality by the operator. In case of movement (resulting in degraded water suppression, degraded water linewidth and/or lipid signal visible in single shots or in accumulated spectrum), the spectral acquisition was stopped (keeping the shots already acquired). Spine T_2_-weighted images were reacquired to reposition the voxel, adjust B_0_ shim again if necessary, and acquire the remaining shots.

### MR Data Processing

#### Available Data

In all, there were 88 visits (20 controls at baseline, 28 FRDA at baseline, 21 FRDA at 1 year, 19 FRDA at 2 years), and 3 datasets were acquired at each visit: Brain anatomical MRI (used for upper cervical spine morphometry), spinal cord dMRI and spinal cord MRS. Out of those 264 datasets, 12 datasets (<5%) could not be used primarily because of excessive motion artifacts, but also artifacts due to braces, or missing data due to subject discomfort or anxiety. The final number of datasets used for the analysis was as follows: Baseline controls: 20 MRI, 18 dMRI, 19 MRS; Baseline FRDA: 26 MRI, 27 dMRI, 26 MRS; 1-year follow-up FRDA: 20 MRI, 20 dMRI, 20 MRS; 2-year follow-up FRDA: 19 MRI, 19 dMRI, 18 MRS.

#### Anatomical MRI

Brain T_1_-weighted images were analyzed using the Spinal Cord Toolbox (SCT) ^40^ version 4.0 to extract spinal cord CSA and eccentricity at C1, C2 and C3 levels. The C2-C3 intervertebral disc was manually delineated to initialize the automatic labeling of vertebrae, after which deep learning algorithms in the SCT toolbox were used to segment the cord.

#### Diffusion MRI

The diffusion images were preprocessed to remove noise ^41^ and Gibbs-ringing artefacts ^42^. The images were then corrected for motion, as well as geometric and eddy-current distortions using FSL (*topup* and *eddy*) ^38^ and bias-field corrected. Subsequently, diffusion tensor fitting using robust estimation of tensors by outliers rejection ^43^ was performed to estimate fractional anisotropy (FA), mean diffusivity (MD), radial diffusivity (RD) and axial diffusivity (AD) ^44^. The mean *b*=0 image was segmented to aid with the registration of the PAM50 template ^45^ into the native diffusion space of each participant. This registration step was initialized using the nonlinear deformation fields generated from the registration of the spine T2-weighted image to the PAM50 template during the segmentation of the spine T2-weighted image. Since it is difficult to identify the vertebral levels in the axially acquired diffusion image, the warping fields between the high-contrast spine T2-weighted image and the template were used to match vertebrae and thus indirectly and automatically label the vertebral levels in the diffusion image. Values for FA, MD, RD and AD within these spinal cord levels were extracted for subsequent statistical analysis.

#### Magnetic Resonance Spectroscopy

Individual shots in the first NT=128 acquisitions were inspected visually and any shot that had severely degraded water suppression, or showed lipid contamination, was removed. In most cases, SNR of metabolites on single shots was not sufficient for shot-to-shot frequency correction, therefore shots were averaged in blocks of 4 or 8 (depending on SNR), then frequency-aligned before summation. The procedure was repeated for the second NT=128 acquisition. The two resulting summed spectra were frequency-aligned and summed to yield a final single spectrum (NT=256). In some participants, one of the two spectra was either not acquired due to time constraints or not usable due to spectral quality, and only one NT=128 spectrum was used in the analysis instead of NT=256.

Spectra were quantified using LCModel 6.3H ^46^. The basis set comprised 18 metabolites: alanine, ascorbate, creatine (Cr), GABA, glucose, glutamine, glutamate, glycerophosphocholine (GPC), glutathione, myo-inositol, scyllo-inositol, lactate, phosphocreatine (PCr), phosphocholine (PCho), phenylethanolamine, N-acetyl-aspartate (NAA), N-acetyl-aspartyl-glutamate (NAAG), taurine as well as an experimentally-measured metabolite-nulled macromolecule spectrum. Metabolites that cannot be distinguished at 3 Tesla are reported as a sum: total N-acetyl-aspartate (tNAA) = NAA + NAAG, total creatine (tCr) = Cr + PCr, total choline (tCho) = PCho + GPC. Based on Cramer-Rao Lower Bounds (CRLB), only 4 metabolites were quantified reliably in the spine (CRLB < 20%): tNAA, tCho, tCr and mIns.

### Effect of Scanner Upgrade

The Trio scanner was upgraded to Prisma in the middle of the study. Out of 21 participants who returned for follow-up, 9 participants were scanned on Trio at baseline and 1-year follow-up, and on Prisma at 2-year follow-up (noted “TTP”), 2 participants were only scanned on Trio (“TT”) (no 2-year follow-up), 7 participants were scanned on Trio at baseline and Prisma thereafter (“TPP”) and 3 participants were scanned only on Prisma (“PPP”). Scanning parameters were matched as closely as possible on Trio and Prisma.

To measure any potential bias due to this scanner upgrade, we scanned 5 additional subjects (not part of the n=20 control participants) on Trio before the upgrade, and on Prisma after the upgrade (see Suppl. Table 1). All measured parameters remained stable before and after the upgrade, with one exception: all diffusivity metrics (AD, RD and MD) were on average 25% higher on Prisma than on Trio. Therefore, in order to mitigate this effect, all underestimated values obtained on Trio were corrected (multiplied) by a factor of 1.25. The reason for this difference was traced to a single mismatched parameter (coil combination mode) set to “Sum of Squares” on Trio, and “Adaptive Combine” on Prisma (“Adaptative Combine” did not exist in VD13D). This resulted in different noise distributions on the two systems ^47^ and affected diffusivity estimates (but not FA). Briefly, the Trio’s “Sum of Squares” method leads to a diffusion signal that follows a noncentral-χ^2^ distribution with an elevated noise floor which artificially increase the diffusion signal, thereby underestimating diffusivity measures. Since we did not keep the raw per-coil data (so-called “TWIX” data), we were unable to reconstruct the Trio data with the “Adaptive Combine” method.

Following this correction of diffusivity values, we performed two analyses of the longitudinal data (which would be the most sensitive to any effect of the scanner upgrade) to rule out any remaining bias: One with all the data, and a second one with “same scanner only” data: For each participant, we kept only time points from the same scanner, i.e. for “TTP” we kept only “TT”, for “TPP” we kept only “PP” and for “PPP” we kept “PPP”. The “same scanner only” longitudinal results were then compared to those obtained with all data (see Suppl. Table 2). We found that longitudinal results were consistent between the two analyses, confirming that longitudinal results from all data were not significantly affected by the scanner upgrade.

### Statistical Analysis

Missing data were left blank. To reduce the extent of multiple testing adjustment, the following metrics were chosen *a priori* (outcome measures):

#### Morphometry

CSA and Eccentricity at C2-C3 obtained from SCT. The average of C2-C3 was chosen to allow comparison with manual segmentation performed at C2-C3 (Suppl. Figure 1).

#### Diffusion

FA, MD, AD, RD averaged over C3-C6 (C2 and C7 were excluded because of inconsistent B0 shimming and image quality at the edge of the field of view).

#### Spectroscopy

Water-referenced concentrations in tNAA, mIns, tCr, tCho, and ratio tNAA/mIns. The ratio tNAA/mIns was chosen because it combines the changes in tNAA (decrease) and mIns (increase) that is observed in many neurodegenerative diseases, maximizing the effect size. For longitudinal slopes and associations between clinical metrics and MR metrics, only the ratio tNAA/mIns was chosen due to higher variability observed in water-referenced concentrations.

#### Clinical metrics

FARS total neuro, SARA, ADL, Functional staging, 9HPT non dominant.

MR metrics in patient and control groups were compared at baseline using linear models with group as the primary variable of interest, and age and sex as covariates.

Among FRDA participants only, associations of clinical metrics (response variables) with MR metrics (predictor variables), using data from all 3 visits, were estimated using linear mixed models, with subjects as random effects.

For each MR metric and each clinical metric, within-participant longitudinal slopes across all visits were estimated using linear regressions. These slopes were tested using one-sided one sample t-tests. For each variable, we hypothesized that longitudinal changes would go in the same direction as cross-sectional differences at baseline. For example, if CSA was lower in FRDA participants than in controls at baseline, we hypothesized that CSA in FRDA participants would continue to decrease over time (null hypothesis: slope < 0). The sample size of the longitudinal cohort (n=21) was sufficient to detect slopes > 0 (or < 0) with an effect size of 0.5 (or -0.5) or better, with α=0.05 and β0.7.

For analyses that were repeated for multiple metrics, reported *p*-values were corrected for type-I error inflation due to multiple testing using the Holm-Bonferroni procedure across metrics within each MR modality (Morphometry: 2 metrics, Diffusion: 4 metrics, Spectroscopy: 5 metrics at baseline, 1 metric longitudinally; Clinical: 5 metrics).

## DATA AVAILABILITY

The data that support the findings of this study are available upon reasonable request from the corresponding author. The data are not publicly available due to their containing information that could compromise the privacy of research participants.

## RESULTS

### Cross-sectional Cohort Findings at Baseline

#### Morphometry

Spinal cord CSA at C2-C3 was 31% lower (p<4.10^−17^) and eccentricity was 10% higher (p<5.10^−7^) in FRDA participants relative to controls (Figure 1 and Table 2). There was no overlap in CSA values between the FRDA and control groups.

**Figure 1.**
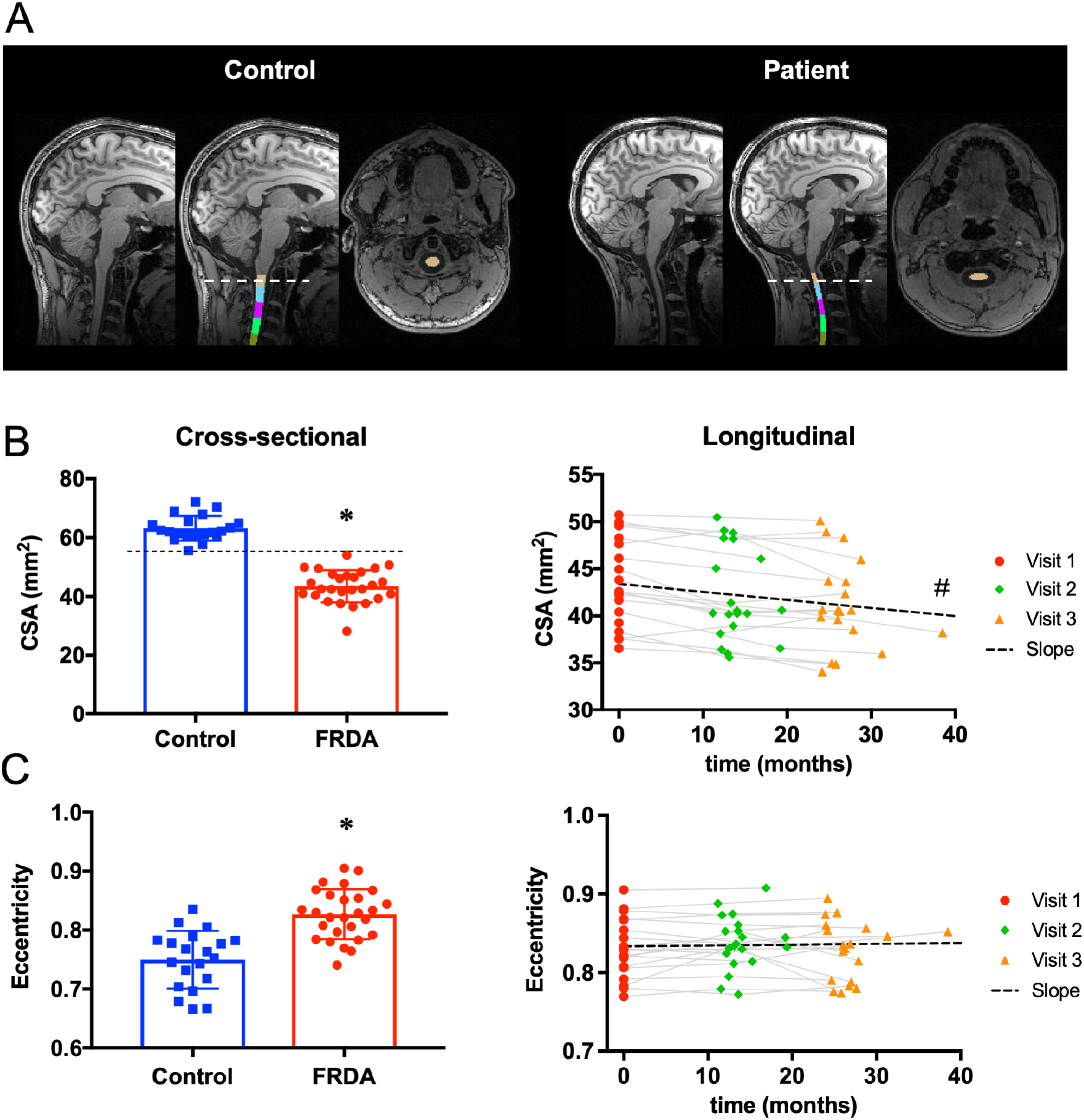
Spinal cord morphometry. **(A)** Brain T1 images showing automated segmentation of the upper cervical spinal cord using SCT. **(B)** Left: Cross-sectional comparison of cross-sectional area at C2-C3 in FRDA vs. Control at baseline. Right: Longitudinal change in cross-sectional area at C2-C3 in FRDA. **(C)** Left: Cross-sectional comparison of eccentricity at C2-C3 in FRDA vs. Control at baseline. Right: Longitudinal change in eccentricity at C2-C3 in FRDA. * FRDA statistically different from control, # slope statistically different from zero.

**Figure 2.**
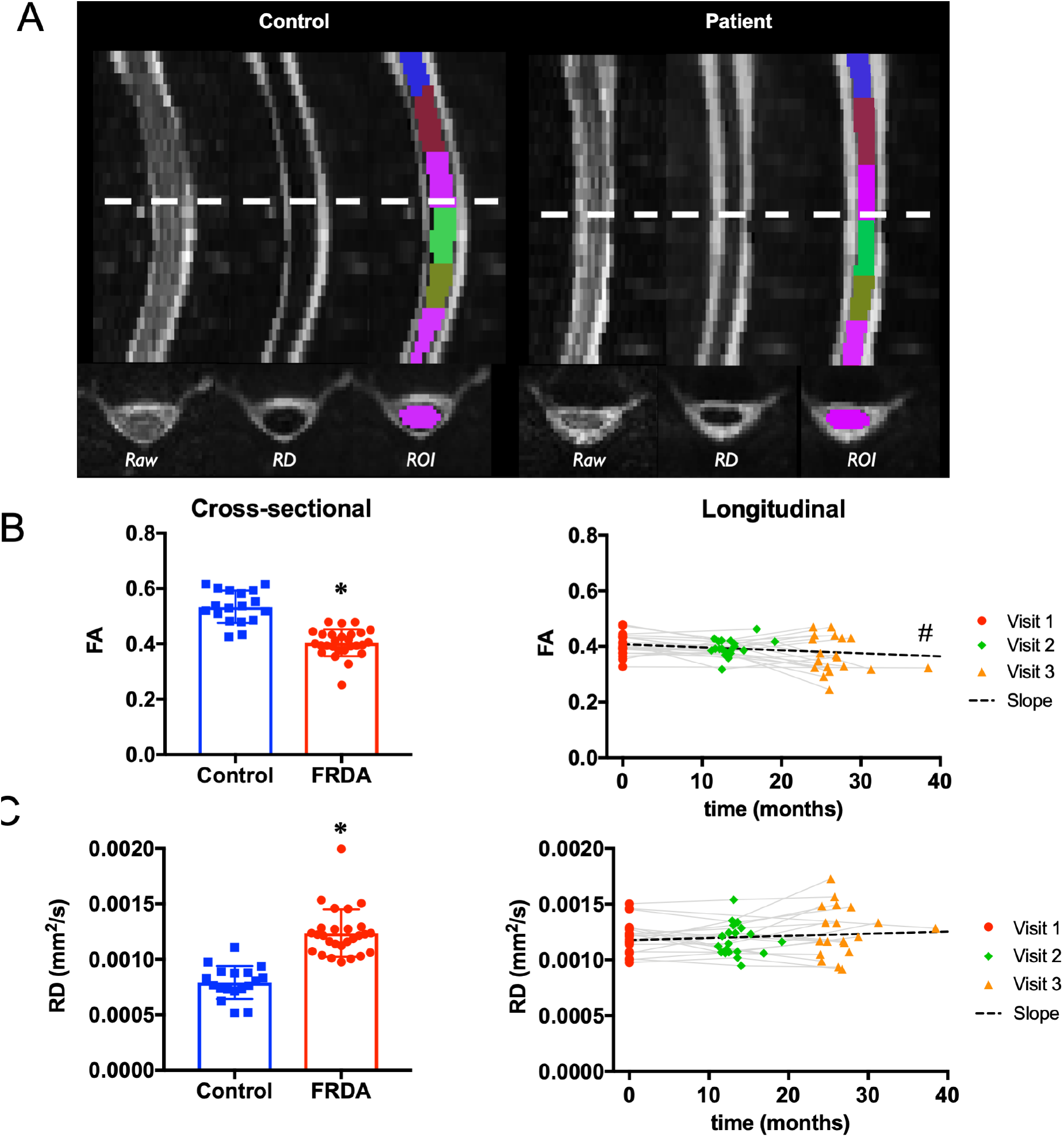
Spinal cord diffusion. **(A)** Diffusion images from a control participant (left), and FRDA participant (right) showing the raw image, the MD image and the ROI segmentation for each participant. **(B)** Left: Cross-sectional comparison of fractional anisotropy (FA) at C2-C3 in FRDA vs. Control at baseline. Right: Longitudinal change in FA at C2-C3 over time in FRDA. **(C)** Left: Cross-sectional comparison of mean diffusivity (MD) at C2-C3 in FRDA vs. Control at baseline. Right: Longitudinal change in MD at C2-C3 over time in FRDA. * FRDA statistically different from control. # slope statistically < 0.

**Table 2.** Cross-sectional differences in the main morphometry, diffusion and spectroscopy parameters at baseline. sd = standard deviation. *** p<0.0005

|  | Modality | CTRL<br>mean (sd) | FRDA<br>mean (sd) | Cohen's d | Difference<br>(%) | Raw p | Corrected p |  |
| --- | --- | --- | --- | --- | --- | --- | --- | --- |
| **CSA (mm<sup>2</sup>) (C2-C3)** | Morphometry | 63.2 (4.2) | 43.7 (5.5) | -3.9 | -31% | 2E-17 | 4E-17 | *** |
| **Eccentricity (C2-C3)** | Morphometry | 0.75 (0.05) | 0.83 (0.04) | 1.7 | 10% | 5E-07 | 5E-07 | *** |
| **FA (C3-C6)** | Diffusion | 0.53 (0.06) | 0.40 (0.05) | -2.5 | -24% | 3E-10 | 1E-09 | *** |
| **MD (C3-C6) (10<sup>-3</sup> mm<sup>2</sup>/s)** | Diffusion | 1.17 (0.18) | 1.58 (0.20) | 2.1 | 35% | 7E-09 | 1E-08 | *** |
| **RD (C3-C6) (10<sup>-3</sup> mm<sup>2</sup>/s)** | Diffusion | 0.79 (0.15) | 1.24 (0.21) | 2.3 | 56% | 6E-10 | 2E-09 | *** |
| **AD (C3-C6) (10<sup>-3</sup> mm<sup>2</sup>/s)** | Diffusion | 1.93 (0.28) | 2.26 (0.19) | 1.4 | 17% | 4E-05 | 4E-05 | *** |
| **tNAA/mIns (C4-C5)** | Spectroscopy | 1.16 (0.23) | 0.56 (0.12) | -3.4 | -52% | 4E-14 | 2E-13 | *** |
| **tNAA (C4-C5) (mM)** | Spectroscopy | 8.6 (1.6) | 5.5 (1.0) | -2.4 | -36% | 1E-09 | 6E-09 | *** |
| **mIns (C4-C5) (mM)** | Spectroscopy | 7.6 (1.4) | 10.4 (1.8) | 1.7 | 37% | 8E-07 | 2E-06 | *** |
| **tCr (C4-C5) (mM)** | Spectroscopy | 4.9 (1.0) | 5.1 (1.4) | 0.2 | 5% | 0.49 | 0.98 | ns |
| **tCho (C4-C5) (mM)** | Spectroscopy | 2.4 (0.5) | 2.3 (0.6) | -0.2 | -5% | 0.53 | 0.98 | ns |

#### Diffusion

FA was 24% lower (p=10^−9^) in FRDA participants relative to controls (Figure 3). All three diffusivity values were higher in FRDA participants, with RD 56% higher (p=2.10^−9^), MD 35% higher (p=10^−8^) and AD 17% higher (p=4.10^−5^) than in controls (Table 2).

**Figure 3.**
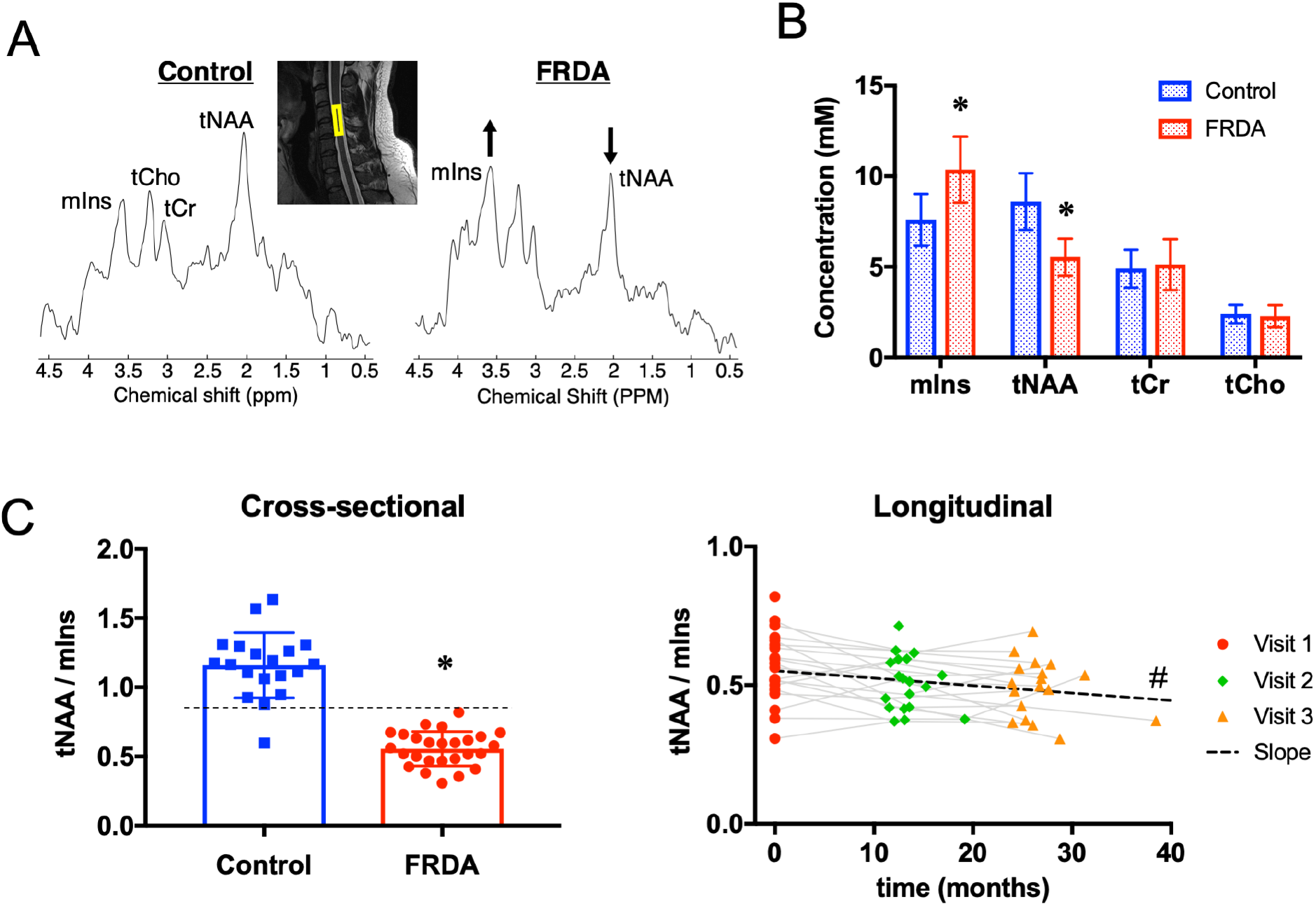
Spinal cord spectroscopy. **(A)** MR spectra from a control participant (left), and FRDA participant (right) clearly showing a lower tNAA peak and a higher mIns peak in FRDA. The inset shows the 8×6×30mm voxel at C4-C5 on a T2 image. **(B)** Cross-sectional comparison of the concentration of the four main metabolites (mIns, tNAA, tCr and tCho) in FRDA vs. Control at baseline. **(C)** Left: Cross-sectional comparison of the ratio tNAA/mIns at C4-C5 in FRDA vs. Control at baseline. Right: Longitudinal change in tNAA/mIns at C4-C5 over time in FRDA. * FRDA statistically different from control, # slope statistically < 0.

#### Spectroscopy

In spite of the small volume, the main resonances (tCho, tCr, tNAA, mIns) were clearly visible in MR spectra (Figure 2). The average water linewidth was 13.0 ± 3.4 Hz, average metabolite linewidth returned by LCModel was 15.4 ± 3.8 Hz and average signal-to-noise ratio (SNR, with noise defined as 2*RMS_noise_) returned by LCModel was 5.0 ± 1.6.

The spectral pattern was markedly different in the FRDA participants relative to controls, with a lower tNAA peak at 2.01ppm and a higher mIns peak at 3.55ppm (Figure 3). The concentration of tNAA was 36% lower (p<6.10^−9^) and the concentration of mIns was 37% higher (p<2.10^−6^) in FRDA participants than in controls (Table 2). As a result, the ratio tNAA/mIns was 52% lower in FRDA participants compared to controls (p<2.10^−13^). There was almost no overlap in tNAA/mIns values between the two groups. All patients had a tNAA/mIns ratio below 0.85, while all controls had a tNAA/mIns ratio higher than 0.85, with one exception: a 10 y.o. control participant had a tNAA/mIns ratio of 0.6.

All cross-sectional results, as well as cross-sectional effect sizes (Cohen’s d), are summarized in Table 2.

### Longitudinal Cohort Findings

Annual rates of change (slopes) for each MR metric and each clinical metric are summarized in Table 3. Effect size is reported as Standardized Response Mean (SRM) defined as mean(slope)/SD(slope).

**Table 3.** Longitudinal annual change (slope) in the main MR and clinical parameters. sd = standard deviation. SRM = Standardized Response Mean. * p<0.05, ** p<0.01, *** p<0.001

|  | Modality | N | Annual slope mean (sd) | SRM | Annual % change from baseline | Raw p | Corrected p |  |
| --- | --- | --- | --- | --- | --- | --- | --- | --- |
| **CSA (C2-C3) (mm<sup>2</sup>)** | Morphometry | 20 | -1.03 (1.09) | -0.95 | -2.4% | 2E-04 | 4E-04 | *** |
| **Eccentricity (C2-C3)** | Morphometry | 20 | 0.0013 (0.0061) | 0.21 | 0.2% | 0.18 | 0.18 | ns |
| **tNAA/mIns** | Spectroscopy | 20 | -0.032 (0.062) | -0.51 | -5.8% | 0.02 | 0.03 | * |
| **FA (C3-C6)** | Diffusion | 20 | -0.013 (0.026) | -0.50 | -3.2% | 0.02 | 0.08 | trend |
| **MD (C3-C6) (10<sup>-3</sup> mm<sup>2</sup>/s)** | Diffusion | 20 | 0.015 (0.085) | 0.18 | 1.0% | 0.22 | 0.44 | ns |
| **RD (C3-C6) (10<sup>-3</sup> mm<sup>2</sup>/s)** | Diffusion | 20 | 0.023 (0.091) | 0.25 | 1.9% | 0.14 | 0.42 | ns |
| **AD (C3-C6) (10<sup>-3</sup> mm<sup>2</sup>/s)** | Diffusion | 20 | -0.007 (0.103) | -0.06 | -0.3% | 0.39 | 0.44 | ns |
| **SARA** | Clinical | 19 | 2.0 (1.1) | 1.89 |  | 8E-08 | 4E-07 | *** |
| **FARS total neuro** | Clinical | 21 | 5.3 (3.7) | 1.44 |  | 1E-06 | 3E-06 | *** |
| **Functional staging** | Clinical | 21 | 0.4 (0.3) | 1.49 |  | 6E-07 | 3E-06 | *** |
| **ADL** | Clinical | 21 | 2.0 (1.4) | 1.39 |  | 2E-06 | 3E-06 | *** |
| **9HPT (non dom.)** | Clinical | 21 | 3.4 (4.0) | 0.86 |  | 4E-04 | 4E-04 | *** |

#### Morphometry

Spinal cord morphometry at C2-C3 using SCT showed a mean rate of atrophy of -2.4% per year (SRM = -0.95), with no significant change in eccentricity (Figure 1). These findings were broadly consistent across individual vertebral levels C1, C2 and C3, with the largest effect size (−1.23) for the average C1-C2 (Suppl. Table 3). The effect size was smaller for C3 (−0.78) than for C1 or C2 (−1.13 and -1.07 respectively) due to higher standard deviation at C3, possibly reflecting the decreasing coil sensitivity and lower SNR at the edge of brain T1 images.

In a separate analysis, spinal cord area and eccentricity were also obtained through manual segmentation of the spinal cord at C2-C3 using SpineSeg ^48^ by three independent raters in a blind fashion (see Suppl. Figure 1). The rate of atrophy from the average CSA across the three raters (−2.9%) was slightly higher than the one found using the automated method SCT (−2.4%) and had slightly lower SRM (−0.87 vs -0.95). All three individual raters found a decrease in CSA over time, but with lower SRM (ranging from -0.62 to -0.79), suggesting that manual segmentation introduces more variability than SCT (Suppl. Figure 1).

#### Diffusion

Fractional anisotropy averaged across C3-C6 showed a trend (raw p<0.02, corrected p<0.08) in yearly decrease of -3.2% with a Cohen’s d of -0.50. There was no significant change for any of the diffusivity metrics (MD, RD, AD) averaged across C3-C6. In post-hoc analyses (Suppl. Table 3), FA values appeared to be most reliable (lower standard deviation and greater effect size) at the center of the imaging volume (C4-C5) were B_0_ shimming is optimal and image quality is higher. FA values averaged between C4 and C5 confirmed the trend in yearly decrease with a change of - 5.6% (raw p<0.0002) and Cohen’s d of -1.0.

#### Spectroscopy

tNAA/mIns showed a significant (p<0.03) yearly decrease of -5.8% with a SRM of -0.51. Because individual metabolite concentrations had higher variability than the ratio tNAA/mIns, slopes for individual metabolites were not included in the main analysis, but are provided in Suppl. Table 3 for information. While none of the individual metabolites showed a statistically significant change over time, numerical results suggest that the longitudinal decrease in tNAA/mIns may be driven both by a decrease in tNAA (−3.9% / year) and an increase in mIns (+0.7% / year). This is supported by the fact that the ratio tNAA/tCr decreased and the ratio mIns/tCr increased, while tCr was relatively stable (Suppl. Table 3).

For comparison, clinical assessments showed an average increase of 2.0 points/year in SARA score (SRM=1.89), 5.3 points/year in FARS total neurological subscale (SRM=1.44), 2.0 points/year in ADL (SRM=1.4) and 0.4 point/year in functional staging (SRM=1.49). The nine-hole peg test (non-dominant hand) showed an average increase of 3.4 seconds per year with an SRM of 0.86.

### Correlations of MR metrics with Clinical Scales

Many of the measured MR parameters correlated with clinical parameters (Table 4). The strongest effects were observed for the spinal cord CSA, followed by FA and tNAA/mIns. CSA correlated negatively with all clinical parameters: FARS, SARA, ADL, functional staging and 9HPT. For example, CSA correlated negatively with FARS total neuro with R=-0.60 and p=4.10^−7^ (Figure 4). FA correlated negatively with FARS total neuro, functional staging and ADL, while tNAA/mIns negatively correlated with functional staging, ADL and 9HPT non-dominant.

**Table 4.** Regression coefficients between clinical parameters and MR parameters. Regressions were performed using data from all 3 time points. Numbers indicate the R coefficient between the corresponding clinical metric (response variable) and MR metric (predictor variable) Blue color indicates negative correlation and red color indicates positive correlation. Higher color intensity indicates higher absolute correlation coefficient * p<0.05, ** p<0.005, *** p<0.0005

|  | SARA | FARS total neuro | Functional staging | ADL | 9HPT (non dom.) |
| --- | --- | --- | --- | --- | --- |
| **CSA (C2-C3)** | -0.55 *** | -0.60 *** | -0.48 *** | -0.51 *** | -0.42 ** |
| **Eccentricity (C2-C3)** | -0.18 ns | -0.25 ns | -0.05 ns | -0.19 ns | -0.14 ns |
| **tNAA/mIns (C4-C5)** | -0.16 ns | 0.02 ns | -0.17 * | -0.31 ** | -0.16 ** |
| **FA (C3-C6)** | -0.26 ns | -0.30 ** | -0.19 * | -0.46 ** | -0.22 ns |
| **MD (C3-C6)** | 0.17 ns | 0.22 ns | 0.12 ns | 0.19 ns | 0.16 ns |
| **RD (C3-C6)** | 0.04 ns | 0.09 ns | 0.00 ns | -0.05 ns | 0.06 ns |
| **AD (C3-C6)** | 0.21 ns | 0.26 ns | 0.17 ns | 0.29 ns | 0.19 ns |

**Figure 4.**
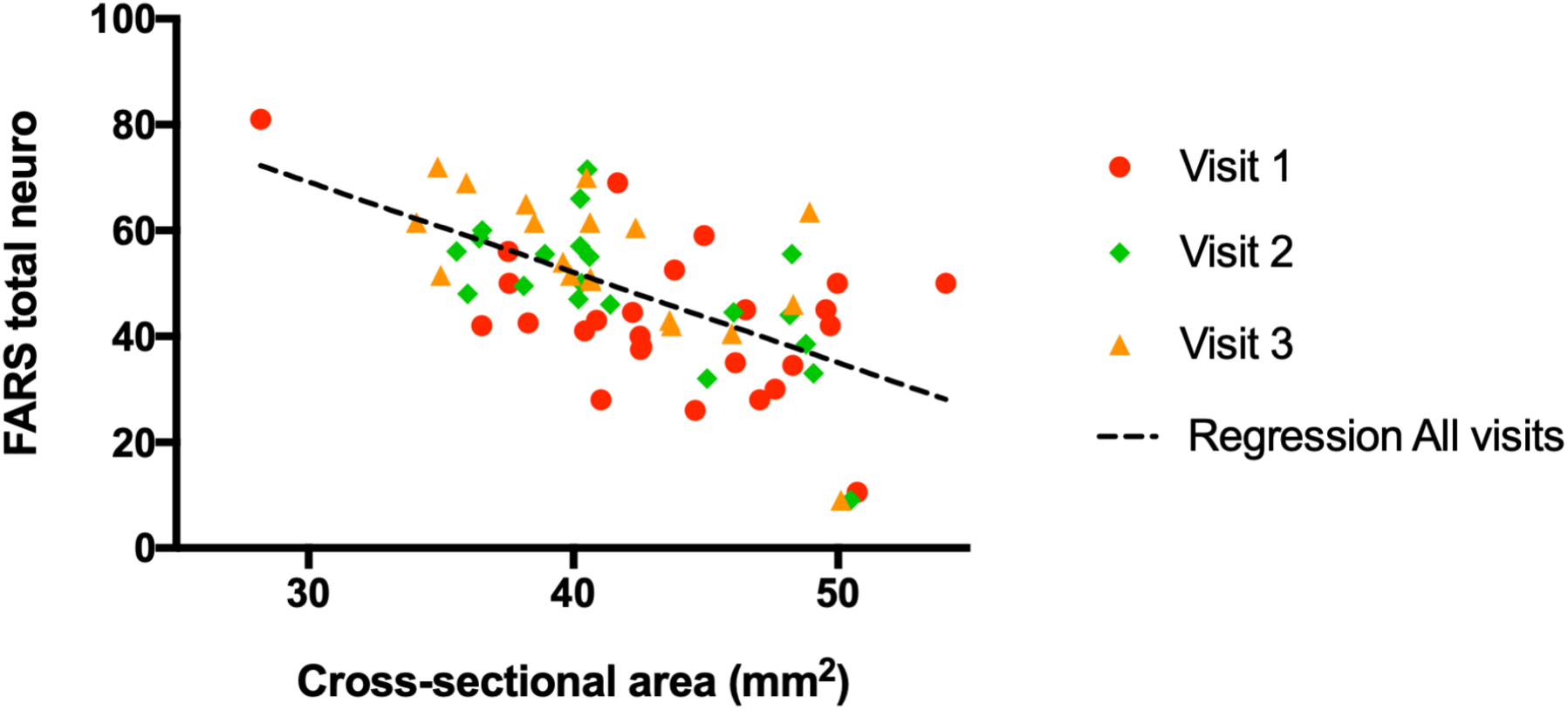
Correlation between FARS total neuro and cross-sectional area. Correlation was determined using cross-sectional area as predictor variable and FARS total neuro as response variable. Data from all three visits were used. R = -0.60, p < 7.10^−6^

## DISCUSSION

To our knowledge, these are the first quantitative *longitudinal* MR results ever reported in the spinal cord in FRDA. Our data demonstrate the ability to monitor disease progression non-invasively in the spinal cord in FRDA, which is critical to assess how potential treatments affect the nervous system in upcoming clinical trials.

In addition, while a handful of studies have reported cross-sectional results in the spinal cord in FRDA with morphometry, and one recent study with DTI but none with MRS, our study provides unique multimodal cross-sectional data. The multimodal approach allowed assessment of anatomy, microstructure and neurochemistry in the same subjects, providing different and complementary measures of neurodegeneration.

Lastly, use of an early-stage cohort (as we did here, with only 5.5 years from onset and 2.3 years from diagnosis) will be crucial for planning of treatment development and testing going forward because efficacy of potential treatments would presumably be maximal if administered in the early stage of the disease, and because the faster disease progression at early stage would make it easier to detect an effect of potential treatments.

All three MR modalities (anatomical MRI, dMRI and MRS) showed large cross-sectional alterations in FRDA participants relative to controls, even at such an early stage. In addition, several MR metrics showed sensitivity to disease progression over time, with spinal cord CSA showing the largest longitudinal effect size among all MR metrics.

### Morphometry

The smaller spinal cord area and higher eccentricity in FRDA compared to controls is consistent with previously reported results using MRI ^11,12,15-17^, as well as post-mortem pathology results ^49^. While these cohorts are not directly comparable, results are remarkably consistent between studies, and show that the upper cervical spinal cord is generally ∼30-40% smaller in FRDA relative to controls, and eccentricity is ∼10% higher.

### Diffusion MRI

Our dMRI cross-sectional findings are consistent with a recent study that reported lower FA and higher diffusivity in total white matter and in specific white matter tracts in FRDA ^30^. Both studies found that FA in total white matter across C3-C6 (C2-C5 in ^30^) was about 25% lower in FRDA participants compared to controls, albeit Hernandez et al. ^30^ reported a smaller cross-sectional effect size (Cohen’s d) of about -0.85 (compared to -2.5 here), which could be due to the larger range of disease severity in their cohort (IQR for FARS total neuro = 41 vs 14.8 in the present study) and therefore larger SD, or due to differences in MR acquisition parameters affecting the SNR of images.

Both studies also found that RD in total white matter was much higher in FRDA than in controls (+56% in our study, about +160% in ^30^), while the increase in AD was smaller (+17% in our study, +6% in ^30^)

The lower FA and higher diffusivity observed in the spinal cord are similar to those observed in the CNS in many neurodegenerative diseases and are consistent with neurodegeneration.

### Magnetic Resonance Spectroscopy

Higher tNAA and lower mIns have been observed in numerous neurodegenerative brain disorders and are not specific to FRDA. However, alterations observed in the spinal cord in the present study were surprisingly large, and much larger than those observed in the brain in FRDA ^50^. In fact, the differences are so pronounced that disease status can practically be determined from visual inspection of the spectra alone.

Myo-inositol is often associated with glia and myelination, while tNAA, synthesized only in neurons, is considered a neuronal marker. Therefore, our findings suggest both neuronal loss and abnormal myelination.

### Longitudinal Effect sizes

Among MR metrics, the largest longitudinal effect size (SRM) was found for cross-sectional area, with an effect size of -0.95 at C2-C3. Post-hoc analyses showed that SRM was even higher for CSA at C1-C2 (−1.23) (Suppl. Table 3). The lower effect size at C3 (−0.78) could be due to lower SNR at the edge of the field of view in the brain T1-weighted image. Studies with larger spinal cord coverage would be needed to investigate longitudinal changes in lower cervical and thoracic areas. However, we hypothesize that C1-C2 will provide close to maximum SRM because that region contains the highest number of ascending axons, and therefore would presumably undergo the largest *absolute* decrease in volume from neurodegeneration.

The longitudinal decrease in FA averaged over C3-C6 was borderline significant after correction for multiple testing, but post-hoc analysis suggests that FA averaged over C4-C5 could be more precise. Indeed, when looking at individual levels (Suppl. Table 3), the SD on diffusion parameters was consistently lowest at C4-C5 and highest at C2 and C7. This suggests that measurement precision is highest in the center of the dMRI field of view, where shim quality is highest.

A tract-specific analysis using SCT, similar to what was recently reported in ^30^, did not reveal improved longitudinal effect size (SRM) in the dorsal columns and corticospinal tract (Suppl. Table 5).

For MRS, the ratio tNAA/mIns was by far the most sensitive longitudinal metric as expected. Other ratios such as tNAA/tCr and mIns/tCr were less sensitive to disease progression, but suggest that both a decrease in tNAA and an increase in mIns contribute to the longitudinal decrease in tNAA/mIns (Suppl. Table 3).

Overall, MR metrics had lower effect sizes than clinical metrics, with spinal cord cross-sectional area coming the closest. However, now knowing where to look, we expect that MR acquisition can be further optimized to improve the effect size of MR metrics. For example, spinal cord CSA was determined here using brain T1 images with 1 mm isotropic resolution and suboptimal SNR below C3 cervical level. Future studies could easily use 0.8mm isotropic, and improved coverage of the cervical cord by activating corresponding coil elements in the 64-channel spine+neck receive coil.

We also noted that SRMs for clinical metrics in the present study appear higher than those reported in literature ^51^. For example, one study reported an increase in FARS total neuro of 4.1 ± 7.84 (Table 10 in ^51^) in a young cohort (BL age < 16) after one year, corresponding to an SRM of 0.52. In the present study, the annual increase in FARS total neuro (from 1-year data only) was 5.2 ± 4.5 corresponding to an SRM of 1.14 (Suppl. Table 4), even though the cohort was older (BL age = 19). It appears that the higher SRM in our study is primarily due to lower SD (4.5 vs 7.84), which could be explained by the fact that all clinical assessments were performed by a single rater (no inter-rater variability).

### True Annual Effect Size

Effect sizes (SRMs) reported in Table 3 are for annual slopes obtained from 2-year data (i.e., 3 time points per participant). To plan clinical trials, it would also be useful to know the “true” annual SRM, namely SRM size obtained from 1-year data only (i.e., 2 time points per participant). These SRMs are reported in Suppl. Table 4. As expected, true annual SRMs were lower than annual SRMs obtained from 2-year data. For example, the effect size for CSA at C2-C3 was -0.95 for annual slope from 2-year data, and -0.71 for annual slopes from 1-year data only.

### Hypoplasia vs. Neurodegeneration

The large differences observed in FRDA participants relative to controls (for example, -52% lower tNAA/mIns) suggest that alterations may already be present well before onset. This is consistent with the hypothesis that alterations in the spinal cord are largely due to hypoplasia rather than neurodegeneration ^49^. While previously reported correlations between CSA and measures of disease severity at baseline suggested that neurodegeneration was also present in FRDA ^15^, our longitudinal data provides direct evidence and quantifies this neurodegeneration, with a rate of atrophy of –2.4% per year in our early-stage cohort.

### MR Metrics for Monitoring Disease Progression in Premanifest Individuals

While CSA was the MR metric with the strongest longitudinal effect size in our cohort, other MR metrics could also play a key role, especially during the pre-manifest phase. For example, it is possible that neurochemical (MRS) and microstructural (DTI) alterations occur earlier during development than alterations in cross-sectional area (morphometry). While identification of premanifest individuals with FRDA is currently rare, perinatal genetic testing will likely become more widespread in the near future once the first effective treatments become available. MR modalities capable of detecting changes during the premanifest phase could therefore play a crucial role in assessing treatment efficacy during that phase, and in determining how early to start treatment.

### Possible Improvements to the MR protocol

With results now providing a better picture of the expected cross-sectional alterations and longitudinal changes in the spinal cord in FRDA, the MR protocol could be streamlined and shortened substantially, benefitting from recent advances in hardware and pulse sequences. For example, while spine dMRI acquisition lasted about 40min in the present study, it can be reduced to ∼4min ^21^. Similarly, the length of spinal cord MRS was about 40min (including all adjustments and shimming). Shorter TR, fewer shots (due to improved SNR with available 64ch head-neck coils), faster shimming using FASTMAP, and improved workflow through automation of adjustment steps now allow acquisition of similar data in about 10min.

### Limitations of the Study

Our study has a number of limitations. First, follow-up scans were only performed in FRDA participants, and not in controls. Therefore, slopes of change over time in FRDA were tested compared to zero, assuming that control values remain stable during the follow-up period. Indeed, published studies suggest that longitudinal changes in control participants are unlikely to explain the changes observed in our FRDA participants (average age 19-20 years, range 10-35 years). For example, large cross-sectional studies in healthy participants across age have shown that CSA increases from age 10 to early-mid-twenties, and remains stable from 20s to 30s ^52,53^. Therefore, we would not expect to see a decrease in CSA in controls in our cohort. For spine MRS, concentrations reported in healthy controls show less than 1% decrease in tNAA and no change in mIns across ages in the 20-70 age range ^54^. For dMRI, FA and MD have been shown to be fairly stable, with less than 0.2% change per year in the 10-40 age range ^55^. Therefore, it seems unlikely that the observed decrease in CSA, tNAA/mIns and DTI metrics in the present study would be due to normal aging.

Another limitation is that the scanner was upgraded midway through the study. However, the test-retest data obtained before and after upgrade, and the fact that we find similar results with “same scanner” data and with “all data” gives us high confidence that the longitudinal changes reported here were not substantially affected by the scanner upgrade.

### Need for Larger, Multi-Site Study

Finally, while our single-site study provides the first quantitative longitudinal results in the spinal cord in FRDA, they were obtained in an early-stage cohort and cannot be generalized to later disease stages. Natural history studies of clinical scales have shown that disease progression in FRDA is faster at early stage ^51^, as well in younger patients and in classical onset FRDA as opposed to late onset ^56^. Our cohort in the present study included only classical onset FRDA participants, most of whom were young and early stage. The average annual increase (slopes from 2-year data) was 5.3 ± 3.7 for FARS total neuro, 2.0 ± 1.1 for SARA and 2.0 ± 1.4 for ADL (Table 3). Based on natural studies of clinical scales, changes in MR metrics are likely slower in older cohorts, late onset cohorts and/or non-ambulatory cohorts.

Larger multi-site studies are needed to cover the entire range of disease severity and duration and gain a fuller picture of disease progression in the spinal cord in FRDA. A new multi-site longitudinal neuroimaging study (TRACK-FA^1^) has now started and aims to scan 200 patients and >100 controls at 3 time points over 24 months. This international effort, involving seven sites on four continents, will use an improved and streamlined neuroimaging protocol that includes the modalities presented in this paper.

## Data Availability

All data produced in the present study are available upon reasonable request to the authors

## Abbreviations

9HPT: 9-hole peg test
AD: axial diffusivity
ADL: activities of daily living
BL: baseline
Cr: creatine
CRLB: Cramer-Rao lower bounds
CSA: cross-sectional area
CSF: cerebrospinal fluid
dMRI: diffusion
MRI EPI: echo-planar imaging
FA: fractional anisotropy
FARS: Friedreich ataxia rating scale
FRDA: Friedreich Ataxia
GPC: glycerophosphocholine
MD: mean diffusivity
mIns: myo-inositol
MRS: magnetic resonance spectroscopy
NAA: N-acetyl-aspartate
NAAG: N-acetyl-aspartyl-glutamate
PCho: phosphocholine
PCr: phosphocreatine
RD: radial diffusivity
SARA: scale for the assessment and rating of ataxia
SCT: spinal cord toolbox
SNR: signal-to-noise ratio
SRM: standardized response mean
tCho: total choline
tCr: total creatine
tNAA: total N-acetyl-aspartate

## Footnotes

1 https://www.monash.edu/medicine/trackfa

