## Supplementary Materials for "Spinal cord MRI and MRS Detect Early-stage Alterations and Disease Progression in Friedreich Ataxia"

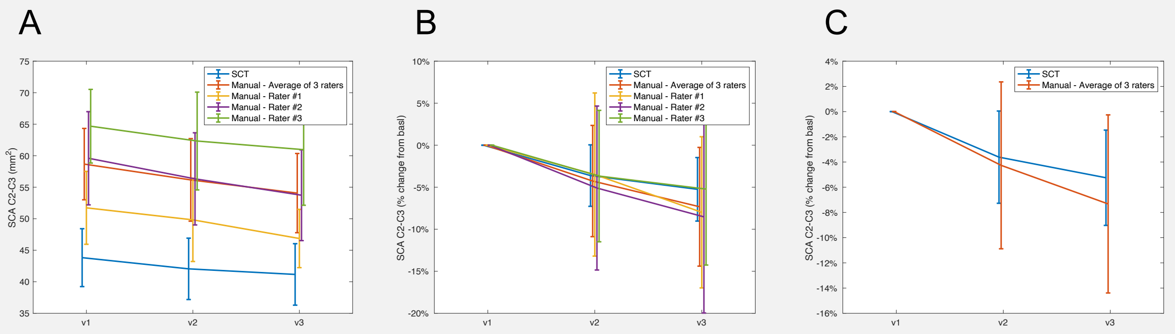


D

| | Mean annual slope | SD annual slope | Annual %change from BL | Effect size (SRM) | Raw p-value |
|---|---|---|---|---|---|
| **CSA (C2-C3) - SCT** | -1.03 | 1.09 | -2.4% | -0.95 | 2E-04 |
| **CSA (C2-C3) - Manual - Average of 3 raters** | -1.66 | 1.92 | -2.9% | -0.87 | 7E-04 |
| **CSA (C2-C3) - Manual - Rater #1** | -1.64 | 2.35 | -3.3% | -0.70 | 3E-03 |
| **CSA (C2-C3) - Manual - Rater #2** | -2.31 | 2.92 | -4.0% | -0.79 | 1E-03 |
| **CSA (C2-C3) - Manual - Rater #3** | -1.57 | 2.55 | -2.5% | -0.62 | 8E-03 |

E

| R | SARA | FARS total neuro | Functional staging | ADL | 9HPT (non dom.) |
|---|---|---|---|---|---|
| **CSA (C2-C3) - SCT** | -0.55 *** | -0.60 *** | -0.48 *** | -0.51 *** | -0.42 ** |
| **CSA (C2-C3) - Manual** | -0.61 *** | -0.62 *** | -0.48 *** | -0.48 *** | -0.42 * |

***Supplementary Figure 1. Comparison of automated segmentation (SCT) with manual segmentation (SpineSeg).*** *SCT results are the same as in the main manuscript. For comparison, the spinal cord was manually segmented by three separate raters who were blind to disease status and visit number. Datasets were segmented in random order. Each rater segmented the cord on three contiguous slices at the C2-C3 intervertebral disk. Values from all three slices were then averaged to yield a single value of CSA and eccentricity per subject and per rater* ***(A)*** *Mean change in cross-sectional area (CSA) across the 3 visits* ***(B)*** *Percent change in CSA relative to baseline.* ***(C)*** *Same as middle graph after removing curves from individual raters for easier visualization.* ***(D)*** *Comparison of longitudinal metrics* ***(E)*** *Comparison of correlation coefficients.*

All 3 raters found a significant CSA decrease over time, ranging from -2.5% to -4.0%. The annual decrease in CSA for the average of all three raters (-2.9%) was comparable to the annual decrease found with SCT (-2.4%). Effect size was smaller for individual raters (ranging from -0.62 to -0.79) than for the average of all 3 raters (-0.87). Effect size for individual raters was also smaller than for SCT (-0.95), suggesting that automated segmentation with SCT is more precise than manual segmentation.

Note that the absolute value of CSA varied significantly across raters as seen in (A). This is because the determination of the CSF/cord boundary is subjective and very dependent on the contrast and brightness of the computer screen used by each rater. However, the % annual change from baseline was remarkably consistent across raters.

**Before correction of diffusivity values**

| | Mean Trio | SD Trio | Mean Prisma | SD Prisma | Diff Prisma vs Trio | Raw p |
|---|---|---|---|---|---|---|
| **CSA (C2-C3) ($mm^2$)** | 70.2 | 7.5 | 70.9 | 7.5 | 0.9% | 0.41 |
| **tNAA/mIns** | 1.03 | 0.16 | 1.13 | 0.23 | 9.3% | 0.2 |
| **FA (C3-C6)** | 0.54 | 0.06 | 0.54 | 0.04 | 0.2% | 0.96 |
| **MD (C3-C6) ($10^{-3}$ $mm^2/s$)** | 0.97 | 0.07 | 1.21 | 0.08 | 25.0% | ***0.002*** |
| **RD (C3-C6) ($10^{-3}$ $mm^2/s$)** | 0.64 | 0.09 | 0.8 | 0.1 | 24.1% | ***0.009*** |
| **AD (C3-C6) ($10^{-3}$ $mm^2/s$)** | 1.61 | 0.11 | 2.03 | 0.09 | 25.7% | ***0.001*** |

**After correction of diffusivity values (multiplication of Trio values by 1.25)**

| | mean Trio | SD Trio | mean Prisma | SD Prisma | Diff Prisma vs Trio | Raw p |
|---|---|---|---|---|---|---|
| **CSA (C2-C3) ($mm^2$)** | 70.2 | 7.5 | 70.9 | 7.5 | 0.9% | 0.41 |
| **tNAA/mIns** | 1.03 | 0.16 | 1.13 | 0.23 | 9.3% | 0.2 |
| **FA (C3-C6)** | 0.54 | 0.06 | 0.54 | 0.04 | 0.2% | 0.96 |
| **MD (C3-C6) ($10^{-3}$ $mm^2/s$)** | 1.21 | 0.09 | 1.21 | 0.08 | 0.0% | 0.99 |
| **RD (C3-C6) ($10^{-3}$ $mm^2/s$)** | 0.8 | 0.11 | 0.8 | 0.1 | -0.7% | 0.88 |
| **AD (C3-C6) ($10^{-3}$ $mm^2/s$)** | 2.02 | 0.13 | 2.03 | 0.09 | 0.5% | 0.84 |

***Supplementary Table 1. Comparison of longitudinal results with "All data" and "Same scanner data".*** *Main MR metrics in five healthy volunteers scanned on Trio (VD13D) before the scanner upgrade and on Prisma (VE11C) after the upgrade. Significant differences were found for diffusivity values. A correction factor of 1.25 was computed as the mean ratio of the values MD, RD and AD on Trio vs Prisma over C3-C6. After correction, there was no significant difference in diffusivity values between Trio and Prisma.*

**All data**

| Subject | v1 | v2 | v3 |
|---|---|---|---|
| #1 | T | T | P |
| #2 | T | T | P |
| #3 | T | T | P |
| #4 | T | T | |
| #5 | T | T | |
| #6 | T | T | P |
| #7 | T | T | P |
| #8 | T | T | P |
| #9 | T | T | P |
| #10 | T | T | P |
| #11 | T | T | P |
| #12 | T | P | P |
| #13 | T | P | P |
| #14 | T | P | P |
| #15 | T | P | P |
| #16 | T | P | P |
| #17 | T | P | P |
| #18 | T | P | P |
| #19 | P | P | P |
| #20 | P | P | P |
| #21 | P | P | P |

**Same scanner data**

| Subject | v1 | v2 | v3 |
|---|---|---|---|
| #1 | T | T | |
| #2 | T | T | |
| #3 | T | T | |
| #4 | T | T | |
| #5 | T | T | |
| #6 | T | T | |
| #7 | T | T | |
| #8 | T | T | |
| #9 | T | T | |
| #10 | T | T | |
| #11 | T | T | |
| #12 | | P | P |
| #13 | | P | P |
| #14 | | P | P |
| #15 | | P | P |
| #16 | | P | P |
| #17 | | P | P |
| #18 | | P | P |
| #19 | P | P | P |
| #20 | P | P | P |
| #21 | P | P | P |

**All data**

| | Mean annual slope | SD annual slope | Effect size (SRM) | Annual %change from BL | Raw p |
|---|---|---|---|---|---|
| CSA (C2-C3) ($mm^2$) | -1.03 | 1.09 | -0.95 | -2.4% | 2E-04 |
| tNAA/mIns | -0.032 | 0.062 | -0.51 | -5.8% | 0.02 |
| FA (C3-C6) | -0.013 | 0.026 | -0.50 | -3.2% | 0.02 |
| MD (C3-C6) ($10^{-3}$ $mm^2/s$) | 0.015 | 0.085 | 0.18 | 1.0% | 0.22 |
| RD (C3-C6) ($10^{-3}$ $mm^2/s$) | 0.023 | 0.091 | 0.25 | 1.9% | 0.14 |
| AD (C3-C6) ($10^{-3}$ $mm^2/s$) | -0.007 | 0.103 | -0.06 | -0.3% | 0.39 |

**Same Scanner data**

| | Mean annual slope | SD annual slope | Effect size (SRM) | Annual %change from BL | Raw p |
|---|---|---|---|---|---|
| CSA (C2-C3) ($mm^2$) | -1.05 | 1.35 | -0.78 | -2.4% | 1E-03 |
| tNAA/mIns | -0.049 | 0.091 | -0.54 | -8.5% | 0.02 |
| FA (C3-C6) | -0.020 | 0.041 | -0.48 | -4.8% | 0.02 |
| MD (C3-C6) ($10^{-3}$ $mm^2/s$) | 0.005 | 0.108 | 0.05 | 0.3% | 0.42 |
| RD (C3-C6) ($10^{-3}$ $mm^2/s$) | 0.025 | 0.118 | 0.21 | 2.1% | 0.18 |
| AD (C3-C6) ($10^{-3}$ $mm^2/s$) | -0.046 | 0.128 | -0.36 | -2.1% | 0.06 |

***Supplementary Table 2. Comparison of longitudinal results with "All data" and "Same scanner data".*** *To confirm that the scanner upgrade did not bias our results, we performed another analysis with "same scanner data". For any given subject, only data from Trio or data from Prisma were used (whichever number of points was highest). The two tables on the left show the data points used with all data and with same scanner data. The two tables on the right show longitudinal results in each case. The all data table is the same as in the main manuscript (Table 3). The same scanner results are consistent with those obtained with all data.*

| | Mean annual slope | SD annual slope | Annual %change from BL | Effect size (SRM) | Raw p |
|---|---|---|---|---|---|
| **CSA (C1)** | -0.96 | 0.85 | -2.2% | -1.13 | 4E-05 |
| **CSA (C2)** | -1.03 | 0.96 | -2.4% | -1.07 | 6E-05 |
| **CSA (C3)** | -1.02 | 1.32 | -2.4% | -0.78 | 1E-03 |
| **CSA (C1-C2)** | -1.00 | 0.81 | -2.3% | -1.23 | 1E-05 |
| **CSA (C2-C3)** | -1.03 | 1.09 | -2.4% | -0.95 | 2E-04 |
| **CSA (C1-C3)** | -1.00 | 0.93 | -2.3% | -1.09 | 6E-05 |

| | Mean annual slope | SD annual slope | Annual %change from BL | Effect size (SRM) | Raw p |
|---|---|---|---|---|---|
| **tNAA/mIns** | -0.03 | 0.06 | -5.8% | -0.5 | 0.02 |
| **tNAA** | -0.21 | 0.88 | -3.9% | -0.2 | 0.15 |
| **mIns** | 0.07 | 1.17 | 0.7% | 0.1 | 0.40 |
| **tCr** | -0.03 | 0.79 | -0.6% | -0.04 | 0.43 |
| **tCho** | -0.03 | 0.37 | -1.3% | -0.1 | 0.36 |
| **tNAA/tCr** | -0.03 | 0.17 | -2.5% | -0.2 | 0.24 |
| **mIns/tCr** | 0.03 | 0.21 | 1.5% | 0.1 | 0.26 |
| **tNAA/tCho** | -0.07 | 0.43 | -2.7% | -0.2 | 0.25 |
| **mIns/tCho** | 0.11 | 0.65 | 2.3% | 0.2 | 0.24 |

| | Mean annual slope | SD annual slope | Annual %change from BL | Effect size (SRM) | Raw p |
|---|---|---|---|---|---|
| **FA (C2)** | -0.042 | 0.131 | -18.1% | -0.3 | 0.1 |
| **FA (C3)** | 0.002 | 0.042 | 0.6% | 0.1 | 0.4 |
| **FA (C4)** | -0.028 | 0.030 | -6.4% | -0.9 | 3E-04 |
| **FA (C5)** | -0.018 | 0.026 | -4.6% | -0.7 | 2E-03 |
| **FA (C6)** | -0.008 | 0.047 | -2.1% | -0.2 | 0.2 |
| **FA (C7)** | -0.005 | 0.062 | -1.5% | -0.1 | 0.4 |
| **FA (C4-C5)** | -0.023 | 0.024 | -5.6% | -1.0 | 2E-04 |
| **FA (C3-C6)** | -0.013 | 0.026 | -3.2% | -0.5 | 0.02 |

| | Mean annual slope | SD annual slope | Annual %change from BL | Effect size (SRM) | Raw p |
|---|---|---|---|---|---|
| **MD (C2)** | -0.136 | 0.474 | -15.8% | -0.3 | 0.1 |
| **MD (C3)** | -0.010 | 0.175 | -0.7% | -0.1 | 0.4 |
| **MD (C4)** | 0.036 | 0.089 | 2.5% | 0.4 | 0.04 |
| **MD (C5)** | 0.002 | 0.102 | 0.1% | 0.0 | 0.5 |
| **MD (C6)** | 0.025 | 0.125 | 1.6% | 0.2 | 0.2 |
| **MD (C7)** | 0.081 | 0.142 | 5.0% | 0.6 | 0.01 |
| **MD (C4-C5)** | 0.024 | 0.082 | 1.6% | 0.3 | 0.1 |
| **MD (C3-C6)** | 0.015 | 0.085 | 1.0% | 0.2 | 0.2 |

| | Mean annual slope | SD annual slope | Annual %change from BL | Effect size (SRM) | Raw p |
|---|---|---|---|---|---|
| **RD (C2)** | -0.107 | 0.367 | -15.9% | -0.3 | 0.1 |
| **RD (C3)** | -0.013 | 0.163 | -1.1% | -0.1 | 0.4 |
| **RD (C4)** | 0.054 | 0.091 | 4.9% | 0.6 | 0.01 |
| **RD (C5)** | 0.025 | 0.100 | 2.1% | 0.2 | 0.1 |
| **RD (C6)** | 0.027 | 0.146 | 2.1% | 0.2 | 0.2 |
| **RD (C7)** | 0.058 | 0.184 | 4.5% | 0.3 | 0.1 |
| **RD (C4-C5)** | 0.039 | 0.085 | 3.4% | 0.5 | 0.03 |
| **RD (C3-C6)** | 0.023 | 0.091 | 1.9% | 0.3 | 0.1 |

| | Mean annual slope | SD annual slope | Annual %change from BL | Effect size (SRM) | Raw p |
|---|---|---|---|---|---|
| **AD (C2)** | -0.193 | 0.692 | -15.7% | -0.3 | 0.1 |
| **AD (C3)** | -0.005 | 0.223 | -0.2% | 0.0 | 0.5 |
| **AD (C4)** | -0.003 | 0.117 | -0.1% | 0.0 | 0.5 |
| **AD (C5)** | -0.039 | 0.134 | -1.8% | -0.3 | 0.1 |
| **AD (C6)** | 0.008 | 0.141 | 0.4% | 0.1 | 0.4 |
| **AD (C7)** | 0.073 | 0.176 | 3.3% | 0.4 | 0.04 |
| **AD (C4-C5)** | -0.014 | 0.103 | -0.7% | -0.1 | 0.3 |
| **AD (C3-C6)** | -0.007 | 0.103 | -0.3% | -0.1 | 0.4 |

***Supplementary Table 3. Annual slopes for cross-sectional area (CSA) and DTI metrics at different levels of the spinal cord, and for MRS for individual metabolites.*** *Annual slopes were obtained by fitting all 2-year data (3 time points per subject: baseline, 1-year and 2-year follow-up). Units for slopes are $mm^2$ for CSA, mM for individual metabolite concentrations and $10^{-3}$ $mm^2/s$ for diffusivities. Metabolite ratios and FA are dimensionless.*

| | mean diff 12mo | SD diff 12mo | SRM diff 12mo | %change from BL 12mo | Raw p |
|---|---|---|---|---|---|
| **CSA (C2-C3) ($mm^2$)** | -0.98 | 1.38 | -0.71 | -2.2% | 6E-05 |
| **tNAA/mIns** | -0.04 | 0.09 | -0.41 | -4.8% | 0.33 |
| **FA (C3-C6)** | -0.02 | 0.05 | -0.35 | -4.0% | 0.01 |
| **MD (C3-C6) ($10^{-3}$ $mm^2/s$)** | 0.018 | 0.123 | 0.15 | 1.4% | 0.02 |
| **RD (C3-C6) ($10^{-3}$ $mm^2/s$)** | 0.029 | 0.127 | 0.22 | 2.5% | 0.19 |
| **AD (C3-C6) ($10^{-3}$ $mm^2/s$)** | -0.009 | 0.167 | -0.05 | -0.1% | 0.09 |
| **SARA** | 2.1 | 1.5 | 1.45 | | 0.37 |
| **FARS total neuro** | 5.2 | 4.5 | 1.14 | | 8E-09 |
| **Functional** | 0.4 | 0.5 | 0.87 | | 5E-09 |
| **ADL** | 1.9 | 1.7 | 1.11 | | 1E-06 |
| **9HPT (non-dom) (s)** | 3.5 | 7.0 | 0.50 | | 9E-09 |

***Supplementary Table 4. 12-month effect sizes (SRM) from 1-year data only.*** *Annual slopes were obtained by fitting 1-year data (2 time points per subject: baseline and 1-year follow-up).*

Cross-sectional

| | | CTRL | FRDA | Difference (%) | Effect size (Cohen's d) | Raw p |
|---|---|---|---|---|---|---|
| Whole cord WM (C3-C6) | FA | 0.53 ± 0.06 | 0.40 ± 0.05 | -24% | -2.6 | 3E-10 |
| | MD ($10^{-3}$ $mm^2/s$) | 1.17 ± 0.18 | 1.58 ± 0.20 | 35% | 2.3 | 7E-09 |
| | RD ($10^{-3}$ $mm^2/s$) | 0.79 ± 0.15 | 1.24 ± 0.21 | 56% | 2.8 | 6E-10 |
| | AD ($10^{-3}$ $mm^2/s$) | 1.93 ± 0.28 | 2.26 ± 0.19 | 17% | 1.3 | 4E-05 |
| Dorsal columns (C3-C6) | FA | 0.61 ± 0.06 | 0.44 ± 0.03 | -28% | -3.7 | 2E-15 |
| | MD ($10^{-3}$ $mm^2/s$) | 1.07 ± 0.13 | 1.47 ± 0.20 | 37% | 2.2 | 1E-08 |
| | RD ($10^{-3}$ $mm^2/s$) | 0.65 ± 0.09 | 1.09 ± 0.16 | 69% | 3.4 | 4E-13 |
| | AD ($10^{-3}$ $mm^2/s$) | 1.92 ± 0.28 | 2.22 ± 0.35 | 15% | 0.9 | 7E-03 |
| Cortico-spinal tract (C3-C6) | FA | 0.55 ± 0.05 | 0.43 ± 0.06 | -22% | -2.4 | 2E-08 |
| | MD ($10^{-3}$ $mm^2/s$) | 1.14 ± 0.16 | 1.48 ± 0.21 | 30% | 2.1 | 6E-07 |
| | RD ($10^{-3}$ $mm^2/s$) | 0.76 ± 0.13 | 1.13 ± 0.23 | 49% | 2.6 | 1E-07 |
| | AD ($10^{-3}$ $mm^2/s$) | 1.91 ± 0.26 | 2.18 ± 0.20 | 14% | 1.1 | 4E-04 |

Longitudinal

| | | Mean annual slope | SD annual slope | Effect size (SRM) | Annual % change from baseline | Raw p |
|---|---|---|---|---|---|---|
| Whole cord WM (C3-C6) | FA | -1E-02 | 3E-02 | -0.50 | -3.2% | 0.02 |
| | MD ($10^{-3}$ $mm^2/s$) | 2E-05 | 9E-05 | 0.18 | 1.0% | 0.22 |
| | RD ($10^{-3}$ $mm^2/s$) | 2E-05 | 9E-05 | 0.25 | 1.9% | 0.14 |
| | AD ($10^{-3}$ $mm^2/s$) | -7E-06 | 1E-04 | -0.06 | -0.3% | 0.39 |
| Dorsal columns (C3-C6) | FA | -9E-03 | 2E-02 | -0.39 | -2.2% | 0.09 |
| | MD ($10^{-3}$ $mm^2/s$) | -2E-05 | 1E-04 | -0.15 | -1.2% | 0.51 |
| | RD ($10^{-3}$ $mm^2/s$) | -2E-06 | 1E-04 | -0.02 | -0.2% | 0.92 |
| | AD ($10^{-3}$ $mm^2/s$) | -5E-05 | 2E-04 | -0.26 | -2.5% | 0.26 |
| Cortico-spinal tract (C3-C6) | FA | -1E-02 | 3E-02 | -0.52 | -3.3% | 0.03 |
| | MD ($10^{-3}$ $mm^2/s$) | 2E-05 | 9E-05 | 0.20 | 1.2% | 0.37 |
| | RD ($10^{-3}$ $mm^2/s$) | 3E-05 | 9E-05 | 0.29 | 2.4% | 0.22 |
| | AD ($10^{-3}$ $mm^2/s$) | -3E-06 | 1E-04 | -0.03 | -0.1% | 0.91 |

***Supplementary. Table 5. Cross-sectional and longitudinal results for two substructures of the spinal cord (dorsal columns (DC) and cortical-spinal track (CST)) compared to whole cord WM.*** *Dorsal columns showed better cross-sectional effect size (Cohen's d) than whole cord WM for FA (-3.7 vs -2.6) and RD (3.4 vs 2.8), but longitudinal effect size and annual change from baseline were more pronounced in whole cord WM. The cortico-spinal tract showed effect sizes and longitudinal changes from baseline comparable to whole cord WM.*
